# Clinical population genetic analysis of variants in the SARS-CoV-2 receptor ACE2

**DOI:** 10.1101/2020.05.27.20115071

**Authors:** Amin Ardeshirdavani, Pooya Zakeri, Amirhosein Mehrtash, Sayed Mostafa Hosseini, Guangdi Li, Hanifeh Mirtavoos-Mahyari, Mohamad javad Soltanpour, Mahmoud Tavallaie, Yves Moreau

## Abstract

**Purpose:** SARS-CoV-2 infects cells via the human Angiotensin-converting enzyme 2 (ACE2) protein. The genetic variation of ACE2 function and expression across populations is still poorly understood. This study aims at better understanding the genetic basis of COVID-19 outcomes by studying association between genetic variation in ACE2 and disease severity in the Iranian population.

**Methods:** We analyzed two large Iranian cohorts and several publicly available human population variant databases to identify novel and previously known ACE2 exonic variants present in the Iranian population and considered those as candidate variants for association between genetic variation and disease severity. We genotyped these variants across three groups of COVID-19 patients with different clinical outcomes (mild disease, severe disease, and death) and evaluated this genetic variation with regard to clinical outcomes.

**Results:** We identified 32 exonic variants present in Iranian cohorts or other public variant databases. Among those, 11 variants are novel and have thus not been described in other populations previously. Following genotyping of these 32 candidate variants, only the synonymous polymorphism (c.2247G>A) was detected across the three groups of COVID-19 patients.

**Conclusion:** Genetic variability of known and novel exonic variants was low among our COVID-19 patients. Our results do not provide support for the hypothesis that exonic variation in ACE2 has a sizeable impact on COVID-19 severity across the Iranian population.

## Introduction

The Coronavirus Disease 2019 (COVID-19) pandemic, which is caused by the Severe Acute Respiratory Syndrome Coronavirus 2 (SARS-CoV-2), continues to spread around the world. At the time of writing of this article, the pandemic had spread to all but a handful of countries worldwide, and the number of COVID-19 cases has sharply risen to 5 million cases, claiming the lives of more than 330,000 people. Iran has one of the highest rates of SARS-CoV-2 infection in the world. As of May 21, 2020, Iran had the tenth-highest number of confirmed cases globally and the ninth-highest number of deaths in the world. With close to 130,000 confirmed COVID-19 cases and more than 7,200 deaths, Iran is currently the country most affected by the pandemic in West Asia.

Four human coronaviruses (HCoV-NL63, HCoV-229E, HCoV-HKU1, and HCoV-OC43) cause respiratory infections with mild symptoms a.k.a. the common cold^1^. However, three other coronaviruses (the Severe Acute Respiratory Syndrome Coronavirus (SARS-CoV), the Middle East Respiratory Syndrome Coronavirus (MERS-CoV), and SARS-CoV-2), which have been transmitted from animals to humans, cause respiratory tract infections that are potentially severe with significant morbidity and mortality^2^. They predominantly affect people with already weak immune systems. While both SARS-CoV and MERS-CoV had higher mortality rates than SARS-CoV2, SARS-CoV2 has already claimed more significantly more lives, possibly because presymptomatic and asymptomic transmission makes epidemic control significantly more difficult^3^.

The Angiotensin-Converting-Enzyme protein 2 (ACE2) was previously identified as the main host cell receptor of SARS-CoV and the human respiratory coronavirus HCoV-NL63^1,4,5^. Recent studies have shown that ACE2 also plays a similar role in the entry of the SARS-CoV2 into the cell^6,7^. The novel SARS-CoV2 binds to membrane-bound ACE2 proteins in their target cells through ACE2 for cellular entry. It has also been reported that ACE2 is mainly expressed by epithelial cells of the lung, intestine, kidney, heart, and blood vessels (it is also lowly expressed in non-vascular cells within the brain)^8^. Previous studies have shown that ACE2 expression is positively correlated with the infection of SARS-CoV and HCoV-NL63 in vitro^9–11^. In particular, it has been confirmed that several human ACE2 variants could decrease the affinity between ACE2 and the spike (S) protein in coronaviruses (HCoV-NL63 and SARS-CoV)^12,13^. Moreover, a recent study provides biophysical and structural evidence that the SARS-CoV-2 spike manifests a 10 to 20 times higher binding affinity to human ACE2 than does the spike of SARS-CoV^14^. This characteristic may also allow the virus to pass more easily from human to human than other β-CoV strains, such as SARS-CoV. Thereby, analyzing allele frequencies (AFs) in different populations, the functional impact of ACE2 gene variants, and gene expression levels in different tissues could provide insight into the variability of disease severity among COVID-19 patients.

As a result, one of the major questions in COVID-19 research today revolves around understanding the role of genetic variation in the pathogenesis of COVID-19. More specifically, which genes and pathways are affected by the underlying genetic variation? Is SARS-CoV2 infection of respiratory epithelia only ACE2-dependent? Do some ACE2 variants provide immunity from infection, or to the contrary increase susceptibility? Do some ACE2 variants contribute to milder or more severe disease progression? More specifically, could some ACE2 variants affect the interaction between the ACE2 and S proteins? Do human ACE2 variants exhibit tissue-specific expression patterns? Are there any ACE2 variants with tissue-specific effects? Is there heterogeneity between patients? Does this ultimately have implications for the treatment of COVID-19?

So far, the genetic basis of ACE2 expression and function in different populations is poorly understood. A recent study in Shanghai investigated and analyzed all putative functional coding-region variants in ACE2 and the expression Quantitative Trait Loci (eQTL) variants among different populations^15^. They did not identify any direct evidence to support the existence of coronavirus S-protein binding-resistant ACE2 variants in some populations^15^. Nonetheless, they highlighted the potential difference in ACE2 expression in different populations and ethnicities in Asia, which in turn may also speculatively play a role in susceptibility to SARS-CoV-2 or disease progression across different populations^15^. These findings indicate that the genomic characteristics of ACE2 in different populations requires further investigation.

This study aims to investigate the genetic basis of the ACE2 expression and function in the Iranian population and address several related and unanswered questions. This includes initially identifying exonic variants in ACE2 and evaluating the allele frequencies (AF) of known variants located in the coding regions of ACE2 in two large Iranian cohorts and also in several publicly available human population variant frequency databases. To investigate the role of genetic variability in the severity of COVID-19 among the Iranian population, we genotype the candidate variants across three groups of patients with different clinical manifestations (mild disease, severe disease, and death) and evaluate their clinical relevance.

## Materials and Methods

To identify exonic variants in ACE2 among the Iranian population, we comprehensively investigate the genomic characteristics of ACE2 in two large cohorts from Iranian genome databases. This gives us insight into the genetic heterogeneity between or within Iranian patient cohorts. To provide a more holistic picture of ACE2 rare and common variant frequencies in the Iranian population, we cross-reference the relevant data in major variant data repositories. All ACE2 gene variants are collected from inhouse and public variant frequencies databases, including GenAP (in-house database of Cimorgh Medical IT Solutions, Iran), Iranome (a catalog of genomic variants in the Iranian population)^16^, 1KGP (1000 Genome Project)^17^, ExAC (Exome Aggregation Consortium)^18^, and gnomAD (Genome Aggregation Database)^19^. We focus on variants that overlap exonic coding regions. ACE2 contains 19 exonic coding regions. We use the coordinates of these variants intersected with coding exons to exclude intronic variants. The constantly up-to-date variant annotation system, eVAI, is used to annotate all variants^20^. AFs for different data sets are provided in Supplementary Table 2. To evaluate variants further, we do not apply any cut-off on AFs because of the limited number of samples available taking into account the total population of Iran (the population of Iran was officially reported to be 81.1 million in 2017) and the significant ethnic diversity of the Iranian population.

We also considered the protein structure of ACE2 and SARS-CoV-2, and investigate the consequences of any of the protein coding variants. To model the structural complex of ACE2 homodimer and the trimeric spike glycoprotein of SARS-CoV-2, two PDB files (6m17, 6vyb) were retrieved from the RCSB protein database^21^. Structural visualization is prepared using PyMOL V2.1^22^.

To assess the relationship between the identified variants and certain clinical aspects of COVID-19 disease, we collected 45 samples from diverse cohorts of individuals with different clinical manifestations. This includes (1) 15 positive COVID-19 individuals (median age 57, range 34–85) with mild symptoms who easily recovered, (2) 15 patients (median age 54, range 36–78) who developed severe symptoms of COVID-19, such as shortness of breath viral, high fever, and pneumonia, but have recovered, and (3) 15 positive individuals (median age 58, range 37–91) who are unfortunately deceased. The detailed information of patients is summarized in Supplementary Table 1.

### Ethics Statement

This study was approved by the Baqiyatallah University of Medical Sciences ethics committee, approval ID: IR.BMSU.REC.1399.041.

## Results

After examining all variants (see Supplementary Table 2) within the exonic coding region of ACE2, we found 32 variants in the Iranian genome variation databases (GenAP and Iranome). Figure 1 shows all of the 32 variants within the coding region of ACE2 (regardless of their effects on the protein structure). 11 of these 32 variants are novel ACE2 variants and have not been reported in the three other databases previously. In particular, we observed that only 3 exonic variants (ACE2: c.2247G>A, ACE2: c.2158A>G, and ACE2: c.2070T>C) in ACE2 identified in GenAP and Iranome also appeared in all aforementioned publicly available variant databases. Detailed analysis of the compared results for all variant databases can be found in the Supplementary Table 2. The ProteinPaint^23^ platform was used to visualize our genome data by mapping the variants onto the ACE2 gene. Cross-checking with the GTEx portal downloaded data did not identify any tissue-specific variant. Checking for the presence of these 32 ACE2 variants in the COSMIC database using the ProteinPaint platform resulted in the following findings: out of 179 unique variants, 3 variants have been identified, namely ACE2: c.2123G>A, ACE2: c.656G>A, and ACE2:c.1741G>T, with these variants being associated with the large intestine, stomach, and central nervous system, respectively.

**Figure 1:**
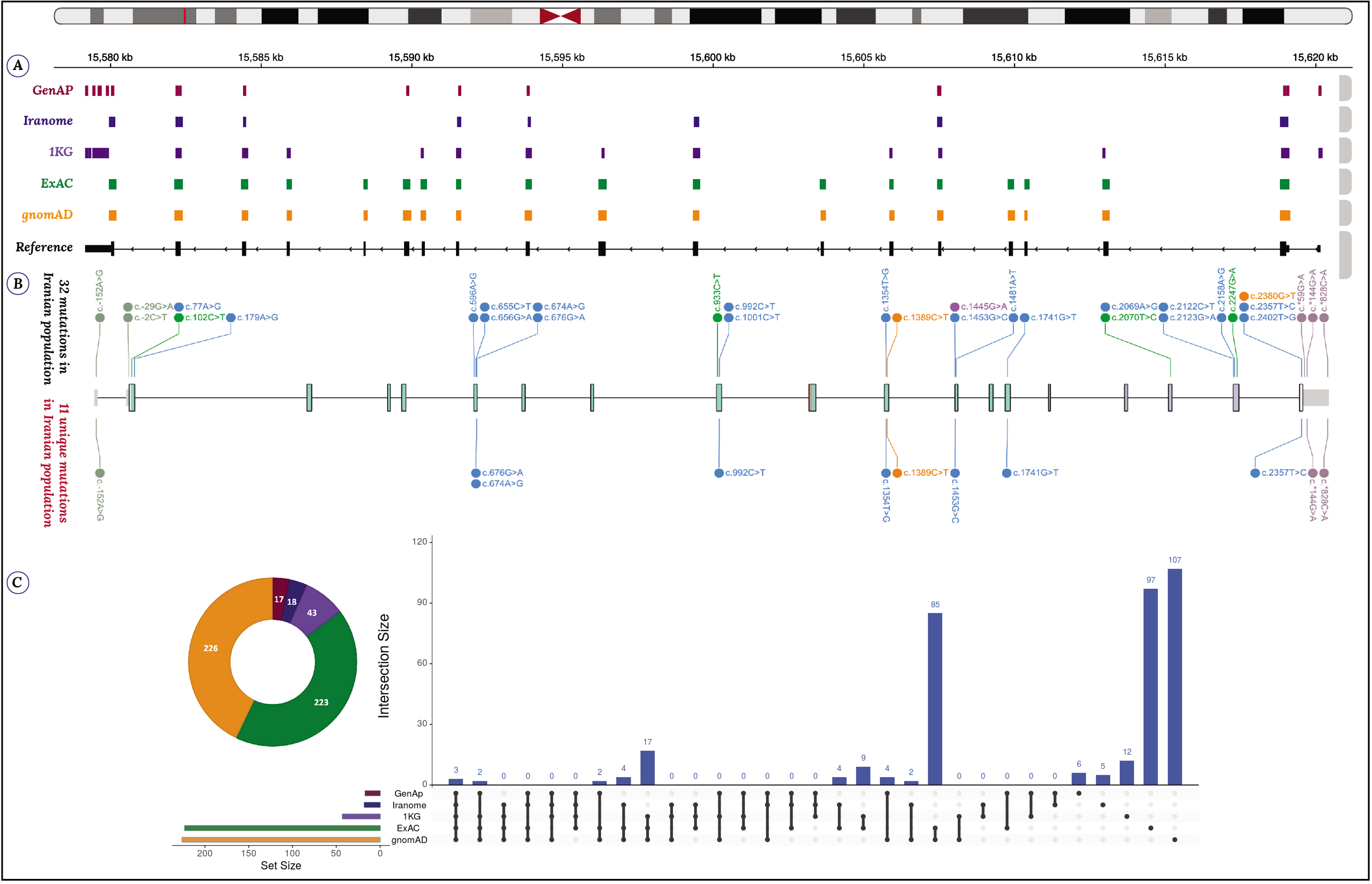
Comparison of five variant databases A) Exonic Variants of five cohorts are mapped onto the human genome reference build CRCh37/hg19 to illustrate their corresponding coverage of the ACE2 gene (IGV^28^). B) The exact location of the 32 variants occurred in the Iranian population. 11 of these 32 variants are novel genetic ACE2 variants and have not been observed in the three other public databases (ProteinPaint^23^). C) The total number of exonic variants found in each database is visualized as a pie chart. The UpSet chart shows the intersections between GenAP, Iranome, 1KG, ExAC, and gnomAD. A closer look at this data can be found in Supplementary Table 2 (Intervene^29^).

Our mapping of exonic variants on the ACE2 protein structure revealed that there are four variants within the ACE2 binding site, namely ACE2: c.1001C>T, ACE2:p.Ser331Phe, ACE2: c.179A>G, and ACE2: c.77A>G. One of these variants (ACE2:p.Ser331Phe) is uniquely present in the Iranian population (Figure 2).

**Figure 2:**
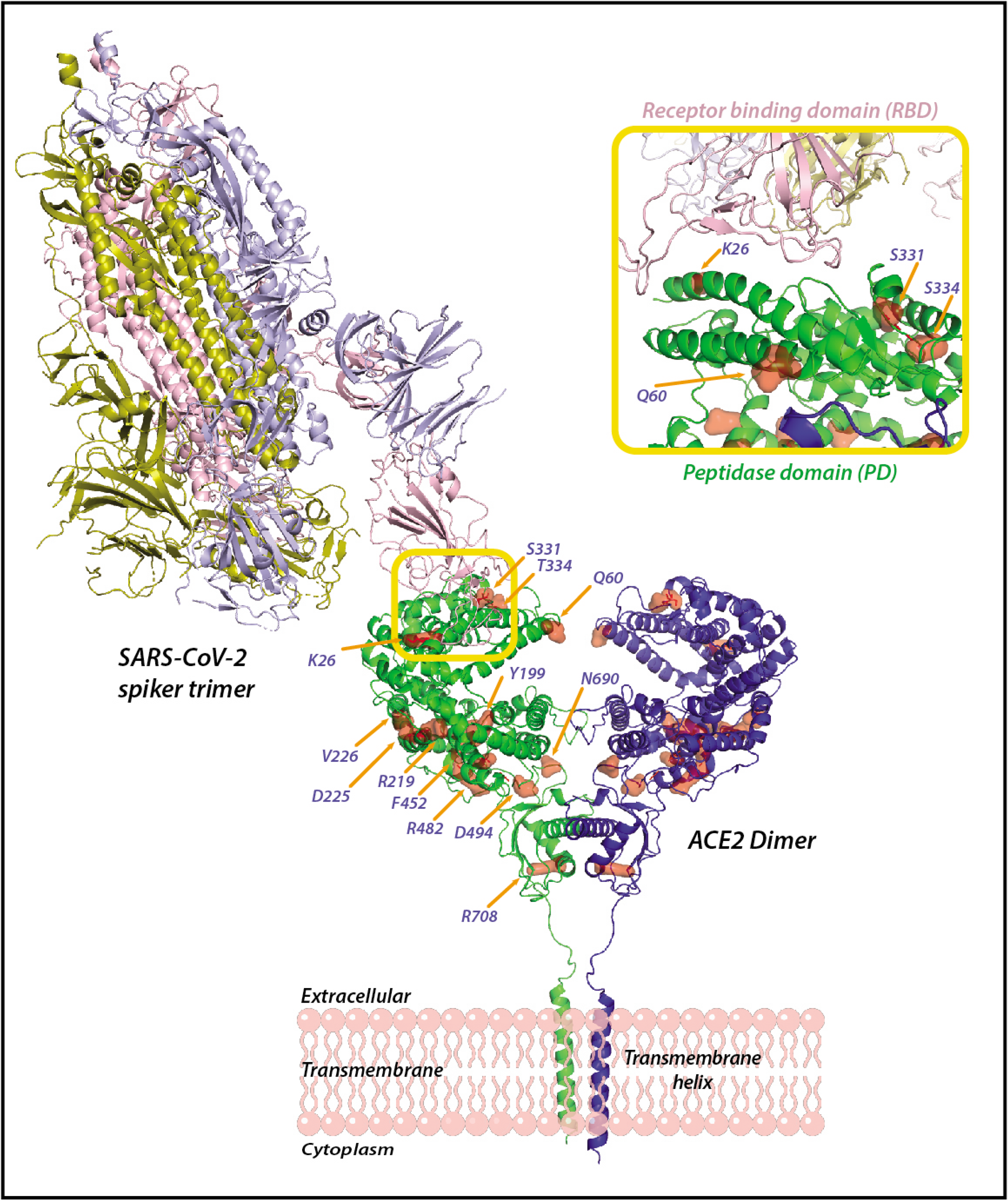
Protein structure of ACE2 and SARS-CoV-2 Substitutions at four amino acid positions, K26, Q60, S331, T334, in the peptidase domain of ACE2 can be in close contact with the receptor-binding domain of the trimeric spike glycoprotein of SARS-CoV-2.

We analyzed the frequency of 32 candidate variants-within the exonic coding region of ACE2 obtained using Sanger sequencing technology-between three groups of patients with different clinical manifestations. In particular, 11 of these 32 genetic ACE2 variants are only reported in Iranian genome data banks: Iranome and GenAP. Table 2 shows that ACE2 exonic variation does not appear to be a key determinant of disease severity or mortaliy among Iranian COVID-19 patients. In fact, in all three groups of patients with different clinical features, no evidence for an association between ACE2 and COVID-19 is detected (see Table 2). In particular, the ACE2 single-nucleotide polymorphism (SNP) K26R –located on exon 2, codon 77 (ACE2:c.77A>G) and common in Caucasians-is not associated with COVID19 clinical outcomes in Iranian patients. A recent study has suggested that the SNP may affect the interaction with SARS-CoV-2 spike glycoprotein and could potentially contribute to the severity of disease^24^.

Moreover, we observed that four individuals –out of forty-five patients considered in this study-carried a variant in exon 18, codon 2247 (ACE2: c.2247G>A –ACE2:c.2247G>A). This included two individuals carrying a heterozygous G-to-A variant and two patients with a homozygous variant (AA). In particular, we found two AG heterozygous individuals and one AA homozygous with mild symptoms. The AA homozygous variant was also observed in a 50-year-old woman with severe symptoms, and was hospitalized but has since recovered. This variant is not seen in patients who did not survive. The ACE2 coding variant, namely ACE2: c.2247G>A, identified in Iranian genome variation databases also appeared in other publicly available variant databases. The variant was associated neither with COVID-19 severity (p = 0.299, Fisher’s exact test) nor with mortality (p = 0.112, Fisher’s exact test). Also, no significant difference is found in the ACE2: c.2247G>A frequencies between individuals with mild symptoms (3 out of 15) and the combined patients of the other two groups (1 out of 29 (p = 0.101, Fisher’s exact test)).

## Discussion

This study provides a comprehensive investigation into the genetic basis of ACE2 function and expression variation in the Iranian population. In summary, our data does not support the hypothesis that exonic variation in ACE2 has a sizeable impact on COVID-19 severity across the Iranian general population. However, these results should be interpreted cautiously because of the small number of patients included in this study, which is a major limitation of this investigation. This limitation, in turn, may lead to potential biases in our analysis. In particular, our results do not preclude that statistically significant effects could be detected for some variants in larger patient cohorts from Iran, although one can speculate that the effect size is likely to be small and thus of limited clinical relevance. Furthermore, our results do not preclude that some of the rare variants identified could have a sizeable effect for individual carriers, although it appears unlikely that rare variants could have a sizeable contribution to the disease burden of the whole Iranian population. Also, we did not assess non-exonic or regulatory ACE2 variation, so we cannot speculate on its potential clinical relevance in the Iranian population. Thus, there is still a need to perform larger clinical studies assessing the role of ACE2 variants in COVID-19 clinical outcomes, in particular for rare variants. Finally, we cannot exclude that exonic ACE2 variation could be relevant in other populations.

While ACE2 is the main host cell receptor of SARS-CoV-2 and plays a central role in the entry of the virus into the cell, it is only one of the many candidate genes that could be associated with the variability of disease severity among COVID-19 patients. Another important candidate is a cellular protease, called Transmembrane protease, serine 2 (TMPRSS2), which appears to be essential for cell entry and the viral spread of the coronavirus^7^. In particular, it has been shown that SARS-CoV-2 employs TMPRSS2 for spike protein priming^7^. Moreover, it has recently been suggested that SARS-CoV-2 may use CD147 –also known as Basigin or EMMPRIN- to gain cell entry^25^. Another potential target is a homolog of ACE2, named Angiotensin-converting enzyme (ACE), which is a major component of the renin-angiotensin system (RAS). ACE2 as a counter-regulatory enzyme of ACE plays an essential role in balancing the activity of ACE in the regulation of blood pressure by contributing to the formation of the RAS axis. ACE is characterized by the existence of a common insertion/deletion (I/D) polymorphism. Several studies have suggested a significant positive correlation between ACE D-allele frequency and disease susceptibility and severity in acute respiratory distress syndrome^26,27^. Accordingly, the assessment of ACE I/D polymorphism alongside clinical outcomes could be helpful to elucidate the variability of disease severity among COVID-19 patients.

## Data Availability

I confirm that all the data related to this study is available.

## Acknowledgments

We are grateful to all patients that participated in this study. We acknowledge the Clinical Research Development Unit of Baqiyatallah Hospital, Tehran, Iran for their guidance and advice. Work by A.A. and Y.M. was supported by Research Council KU Leuven: C14/18/092 SymBioSys3; CELSA-HIDUCTION CELSA/17/032. Flemish Government: IWT: Exaptation, Ph.D. grants. FWO 06260 (Iterative and multi-level methods for Bayesian multirelational factorization with features); Elixir I002819N. This research received funding from the Flemish Government (AI Research Program). VLAIO PM: Augmenting Therapeutic Effectiveness through Novel Analytics. EU: “MELLODDY” This project has received funding from the Innovative Medicines Initiative 2 Joint Undertaking under grant agreement No 831472. This Joint Undertaking receives support from the European Union’s Horizon 2020 research and innovation programme and EFPIA. We would like to thank Prof. Bart De Strooper for the invaluable support he provided to P.Z. during this research.

## Author Contributions

A.A., P.Z., and Y.M. organized, prepared, and wrote the first draft of the manuscript. A.A. conceived and constructed the study, he also collected and developed data sources. A.A., and P.Z. designed the evaluation procedures and analyzed the results. A.M. did primer-design and Sanger sequence-data analysis. S.M.H., M.J.S., and M.T. were in charge of sample collection and sequencing. P.Z. and H.M.M. did the clinical and biological assessment of the study and results. G.L. analyzed and visualized the protein structure. A.A., M.T., and Y.M. supervised the study.

